# Repeatability of a Dual-Scheimpflug Placido Disc Corneal Tomographer/Topographer in Eyes with Keratoconus

**DOI:** 10.1101/2020.05.13.20067710

**Authors:** Anne Poulsen, Diane Jang, Mahmood Khan, Zaina Nabil Al-Mohtaseb, Michael Chen, Xiang Zhan, Kalins Banerjee, Ingrid U Scott, Seth M Pantanelli

## Abstract

**Purpose:** To investigate the repeatability of a combined Dual-Scheimpflug placido disc corneal topographer (Zeimer Galilei G4) with respect to keratometric indices used to monitor progression of keratoconus (KCN).

**Methods:** Patients with KCN were prospectively enrolled. For each eye lacking history of corneal surgery, 5 measurements were taken in succession. Eyes in which 3 or more measurements could be obtained (defined by the device’s 4 image quality metrics) were included in the analysis. The repeatability limits (RL) and interclass correlation coefficients (ICC) were calculated for various parameters.

**Results:** 32 eyes from 25 patients met all image quality metrics, and 54 eyes from 38 patients met at least 3/4 criteria (all except the placido image quality metric). RLs for key parameters when 4/4 or ≥3/4 image quality metrics were met included: 0.37 and 0.77 diopters (D) for steep simulated keratometry, 0.79 and 1.65 D for maximum keratometry, 13.80 and 13.88 degrees for astigmatism axis, 0.64 and 0.56 μm for vertical coma magnitude, and 3.76 and 3.84 μm for thinnest pachymetry, respectively. The ICCs for all parameters were excellent [above 0.87 except for spherical aberration (0.77)].

**Conclusions:** The dual-Scheimpflug placido disc corneal topographer is highly repeatable in quantifying parameters used in monitoring KCN. Excellent placido images are difficult to capture in eyes with KCN, but when available, increase the reliability of the measurements. The RLs may be especially helpful in detecting progression in mild KCN when interventions such as corneal cross-linking or intrastromal corneal ring segments are most beneficial.

## Introduction

Keratoconus is the most common corneal degeneration, with an estimated prevalence of 54.5 per 100,000 in the general population^1^. The degeneration results in significant visual impairment secondary to irregular astigmatism and high myopia. In the age of corneal cross-linking, early diagnosis and detection of progression are essential elements of management that limit morbidity associated with the disease. Achieving a consensus in the definition of progression in keratoconus is limited by the requirements that such indices be based upon high quality data (which is often difficult to obtain in keratoconic corneas) and long-term follow-up in eyes that progress but do not have a surgical intervention (which may be impractical or unethical). One recent study by Gupta et al evaluated the change in several keratometric indices over time and determined that variation in best fit sphere radius was one of the most sensitive predictors of progression^2^. Another study by Martinez-Abad followed patients with keratoconus for 6 years and proposed a “Progression Index” equation, but this study was limited in that it was based only upon anterior surface indices^3^.

The Global Consensus on Keratoconus and Ectatic Disease defined ectatic progression by a consistent change, where the magnitude of the change is above the noise of the testing system^4^. In conforming to these guidelines, a study was published by de Luis Eguileor et al, which investigated the repeatability of measurements pertinent to the monitoring of progression in eyes with keratoconus using a Scheimpflug system (Pentacam HR)^5^. This study found that sequential measurements differing by more than 1.54 D, 1.08 D, and 0.71 ¼m in the steep keratometry, maximum keratometry, and vertical coma, respectively, likely represented real change. Of note, this study only included eyes for which high quality data were available in the analysis. The study results are interesting and immediately applicable for eye care providers who have a Pentacam HR available. However, at least two previous studies have shown that the measured indices from one device are not interchangeable with those of another, like the Orbscan II (Bausch & Lomb, Rochester, NY, USA) or Galilei Dual Scheimpflug Analyzer (Zeimer, Port, Switzerland)^6,7^. It is therefore unclear how these repeatability limits reported by de Luis Eguileor et al would translate for use with a different device, particularly a device employing placido-based or a combination of keratometric technologies. Furthermore, it is unclear whether such repeatability limits can be applied in the setting of sub-optimal image quality, which commonly occurs when measuring keratoconic eyes.

The objective of the current study was to assess the repeatability of measurements obtained using the Galilei Dual Scheimpflug Analyzer (Zeimer, Port, Switzerland) and, thus, to determine which of these measurements have repeatability limits that may allow accurate monitoring of keratoconus progression over time. Since the Galilei captures placido images and the Pentacam HR does not, we also sought to investigate the contribution good placido images have on reliability of these measurements.

## Methods

We conducted a prospective observational study of patients seen at either the Penn State Eye Center (Penn State College of Medicine, Hershey, PA) or the Cullen Eye Institute (Baylor College of Medicine, Houston, TX). The study was approved by the Institutional Review Board at both academic institutions. Patients with keratoconus being seen in routine follow-up were provided with information about the study and written signed consent was obtained prior to inclusion. All keratoconus patients over the age of 18 were offered the opportunity to participate. A diagnosis of keratoconus was made with slit lamp examination and topographic confirmation, and met criteria for at least Stage I Amsler-Kremeich classification^8^. Exclusion criteria included history of any corneal or intra-ocular surgery (i.e. corneal cross linking, LASIK, corneal transplant, or cataract surgery). If a surgery had been performed in one eye, the fellow eye was still eligible for inclusion.

The Galilei G4 is a combined dual-Scheimpflug tomographer / placido disc topographer that employs monochromatic light sources at 470 nm and 750 nm for Scheimpflug and placido disc illumination, respectively. Each scan generates over 122,000 data points. The placido and Scheimpflug data are integrated using a proprietary algorithm to generate information about the anterior surface of the cornea, while only the Scheimpflug data are used to generate information about the posterior surface.

Before starting the study, the Galilei G4 was calibrated at both institutions. Each eye was aligned to the visual axis via the fixation light on the machine. Patients were instructed to blink between measurements. Five consecutive measurements were obtained in each eligible eye, using the standard resolution setting. All images for an individual patient were captured by a single examiner, but given the multi-center nature of this study, multiple examiners were used to obtain data from different patients. The instrument defines 4 image quality metrics to qualify the integrity of the data collected. Satisfactory metrics include a motion compensation value > 85%, placido image quality > 85%, Scheimpflug image quality > 90%, and motion distance > 70%.

For each captured image, the following keratometric indices were assessed: flat and steep simulated keratometry (SimK) magnitudes, steep SimK axis, maximum keratometry values in the central zone (Kmax), anterior (BFS_anterior) and posterior (BFS_posterior) radius best fit sphere of the cornea with the diameter set to 8 mm, maximum anterior (max_BFS_ant) and posterior elevations (max_BFS_post), area of thinnest pachymetry (TCT), corneal asphericity (Q) at 6 mm, spherical aberration 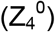, vertical 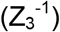 and horizontal 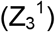 coma, coma axis, root-mean-square (RMS) of coma measurements, and root-mean-square of higher order aberrations (HOA RMS) at 6 mm.

Repeatability limits (RL) for the above indices were calculated, and represent the value over which a change is likely to be real and not due to noise or variability inherent to the machine^9^. The intraclass correlation coefficient (ICC) was also calculated for each index, which estimates the percentage of the total variability in the results that can be attributed to real differences between measurements. The RLs and ICCs were calculated for two different conditions: 1) eyes with at least 3 measurements that met all four of the devices image quality metrics and 2) eyes with at least 3 measurements that met 3 or more of the devices 4 image quality metrics (an excellent placido image was not required). In the instance where 4 or 5 high quality images were obtainable, all of the available data was used in the analysis. Statistical analyses were carried out using R programming language.

## Results

Eighty-six eyes from 51 patients were measured; 32 eyes from 25 patients met all four image quality metric criteria and 54 eyes from 38 patients met at least 3 of 4 criteria. The severity of keratoconus for eyes included in each of the analyses is detailed in Table 1. The RLs and ICCs for all parameters are shown in Table 2. We found that obtaining measurements that met all four image quality metrics was challenging - the placido image criterion was the most difficult to meet. Dropping the requirement for a good placido image and forcing the device to rely more heavily upon the Scheimpflug data lead to higher (worse) RLs. The ICC for all parameters was found to be excellent (above 0.87) except for spherical aberration (0.77).

For the 54 eyes that had less than 3 images captured that met all four of the image quality metrics, the Kmax was 56.02 ± 6.44. Of these, all 54 had poor quality placido images and 27 also had poor Scheimpflug images. An inverse relationship with moderately strong correlation was found between Kmax and the device’s interpretation of the placido (Figure 1a; R^2^ = 0.68), but not Scheimpflug (Figure 1b; R^2^ = 0.08) image quality. This suggests that the placido image was more sensitive to degradation from corneal steepening in moderate or advanced keratoconic disease.

**Figure 1.**
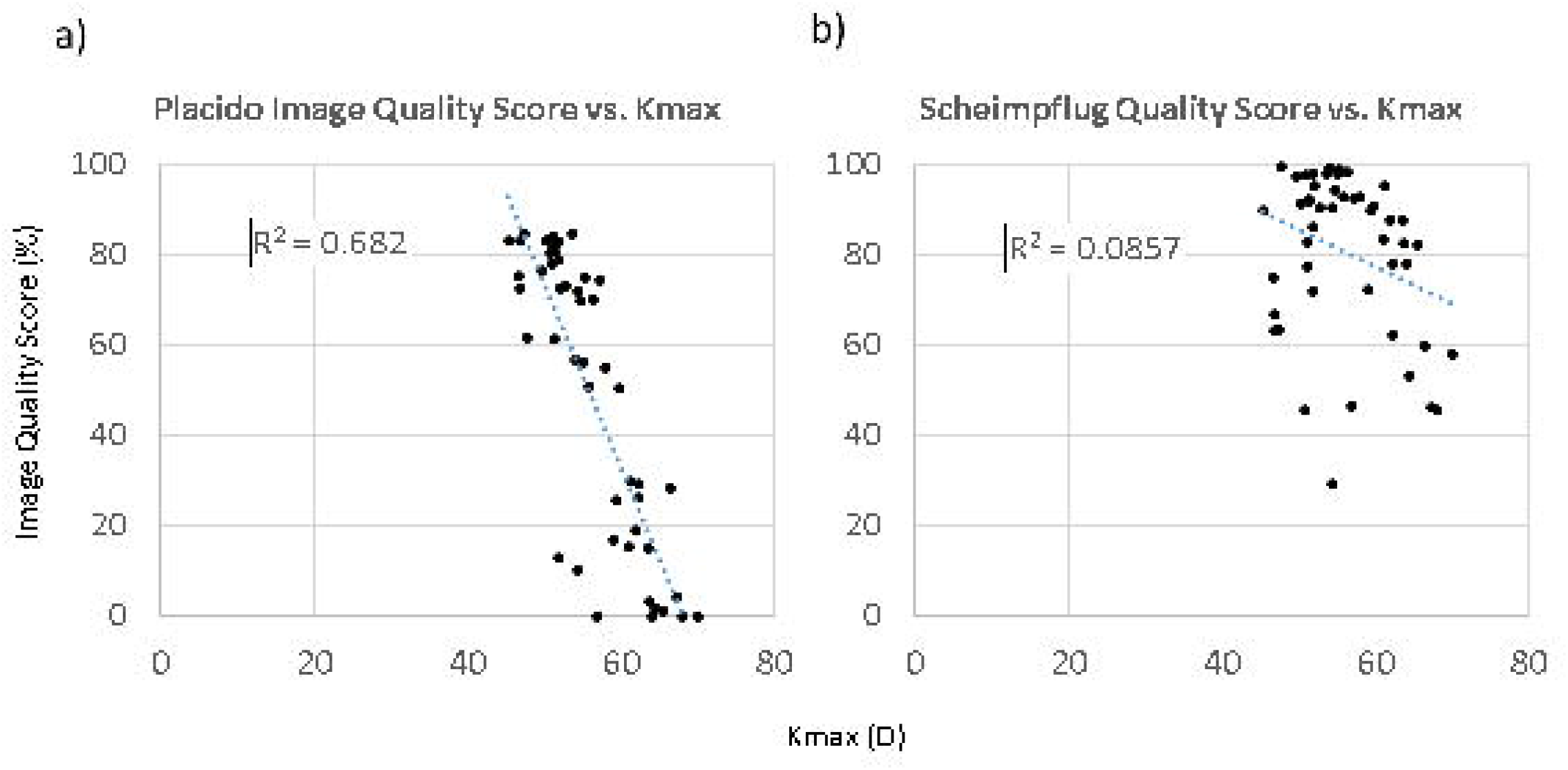
- Placido (a) and Scheimpflug (b) image quality scores (%) as a function of Kmax (D). There was a strong correlation between increasing maximum keratometry and degradation of the placido image, but not the Scheimpflug image.

## Discussion

There are at least two previous studies that have investigated the repeatability of the Galilei in a keratoconic population. Previously, Shetty et al evaluated 55 keratoconic eyes on three different devices and found that the Galilei was the least repeatable when compared to the Pentacam HR and the Sirius (Costruzione Strumenti Oftalmici, Florence, Italy)^7^. However, these investigators only compared performance with respect to three keratometric indices (anterior keratometry, thinnest corneal thickness, and posterior keratometry). More recently, Meyer et al evaluated 50 keratoconic eyes and found the Pentacam to be superior to the Galilei with respect to keratometry measurements, but found the reverse to be true with respect to pachymetry measurements^6^. It is important to note, however, that all eyes, including those for which high quality measurements could not be obtained, were included in the Meyer study analysis, and that 25% of eyes on the Galilei and 12% on the Pentacam HR were reported to have poor quality scans. The rationale for including these data is understandable; especially in abnormal eyes, high quality measurements may not be obtainable. However, the disparate proportions of poor quality data included in the analysis could also skew the results in favor of one device over the other. It is well understood that poor quality data leads to less repeatable measurements, but the present study is the first to quantify how much these repeatability limits are influenced by image quality.

In the present study, RLs and ICCs were excellent when all four of the image quality metrics were met. However, when a good placido image was not necessarily available, the device was still able to use available Scheimpflug data to reliably report on various keratometric indices. Dropping the requirement for a good placido image appeared to affect the RLs for the magnitude of various keratometric indices more than it did the axes. For example, the RLs for steep keratometry in the central zone (SimK; 0.37 and 0.77 D), maximum keratometry (Kmax; 0.79 and 1.65 D), and maximum anterior elevation (max_BFS_anterior; 3.66 and 7.17 μm) all worsened when dropping the requirement for a good placido image; in contrast, RLs for steep keratometry axis in the central zone (SimK axis; 13.80 and 13.88 degrees) and coma axis (32.65 and 25.62 degrees) remained relatively unchanged. We therefore propose that poor placido images do not warrant dismissal of the measurement altogether, assuming Scheimpflug and other image quality metrics are met. The data must just be interpreted in a different context.

Many of the RLs found in the present study were lower than those in the de Luis Eguileor study^5^. This was especially true when all four of the devices image quality metrics were met. We hypothesize that this is due to the Pentacam HR’s reliance upon a single Scheimpflug camera. In contrast, the Galilei G4 employs placido disc video keratography and combines it with two Scheimpflug cameras. Having two Scheimpflug cameras might also make the Galilei less susceptible to motion artifact and parallax, since every point on the cornea is being viewed from two different perspectives at any given time. Alternatively, the differences in RLs and ICCs reported in the de Luis Eguileor study and current study may be due to differences in the severity of keratoconus in the two study populations. A relationship between keratoconus severity and measurement reliability was previously investigated by Hashemi et al, who found that reliability of measurements was acceptable for all tested devices when mild keratoconics were considered, but that reliability dropped off significantly when the maximum K reading was greater than 55.0 D^10^. Many moderate and severe keratoconic eyes were imaged in the present study, but were excluded from the analysis due to poor image quality. It would follow that inclusion of these eyes would have increased RLs and ICCs.

The Global Consensus on Keratoconus and Ectatic Disease previously defined ectatic progression by a consistent change in 2 of the 3 following parameters, where the magnitude of the change is above the noise of the testing system: 1) steepening of the anterior corneal surface, 2) steepening of the posterior corneal surface, or 3) thinning and/or an increase in the rate of corneal thickness change from the periphery to the thinnest point^4^. With regards to steepening of the anterior corneal surface, multiple authors have suggested a magnitude of change in Kmax greater than 1.0 D as indicative of progression^11-13^. Since the present study found RLs for Steep SimK, Kmax, and BFS_anterior of 0.37 D, 0.79 D, and 0.04 mm (~ 0.25 D change from mean BFS_anterior), respectively, we conclude that these three parameters may be used to diagnose progression with the tested device. Similarly, the RL for the BFS_posterior was 0.06 (~ 0.5 D change from mean BFS_posterior), and thus may also be a valid metric used to satisfy the second criterion. With regards to thinning, Vinciguerra and Caporossi suggested a change in thinnest corneal thickness (TCT) of 20 and 10 μm, respectively, as a criterion for defining progression after cross-linking^14,15^. Since the present study’s RL for this metric was 3.76 μm, this metric can be used to satisfy the third criterion. It should be emphasized that these metrics likely cannot be applied to poor quality images or those from moderate and severe keratoconus, where RLs are likely higher.

The present study results also have implications with regards to the implantation of ICRS. Alfonso et al previously proposed criteria for ICRS implantation which included 1) a difference between the flat axis of the corneal cylinder measured with a Javal keratometer and with an Orbscan IIz of less than 30 degrees and 2) a coma axis within 30 degrees of the flat topographic axis^16^. The RL for the coma axis in the present study was high at 32.65 degrees, suggesting that this metric cannot be relied upon to satisfy the Alfonso criteria when planning ICRS orientation.

A limitations of this study includes the relatively small sample size, though similar to that of previously published works in the same domain^6,7^. Secondly, many eyes with clinically moderate or severe keratoconus were scanned but not included in the data analysis since neither good placido nor Scheimpflug images were unobtainable. As such, the results of this study are most applicable to eyes with Stage I and II Amsler-Krumeich keratoconus. However, detection of progression is also most important in this sub-population, in whom consideration for ICRS or corneal cross-linking is most often applicable. It is much less important to know whether progression is occurring in an eye with Stage III or IV keratoconus, where the only option remaining for vision restoration is cornea transplant surgery.

In conclusion, we found that the Galilei Dual Scheimpflug Analyzer was highly repeatable in quantifying parameters typically used in monitoring for keratoconus. Parameters with especially low RLs included Steep SimK, anterior and posterior best fit sphere radius of curvatures, and thinnest corneal thickness. Axis measurements and aberrometric parameters (i.e. coma and spherical aberration) were less reliable, but may still be used to detect progression when interpreted in the context of high image quality and milder keratoconic disease. While the placido videokeratography information likely aids in the Galilei’s sensitivity to keratometric indices, it is also responsible for quickly degrading the quality of the data in more advanced disease states. Poor quality placido images do not warrant dismissal of the measurement altogether, assuming Scheimpflug and other image quality metrics are met. The data must just be interpreted in the context of more relaxed repeatability limits.

## Data Availability

Full dataset may be available to interested parties by request. Please e-mail the corresponding author to query.

## Acknowledgements

The authors would like to acknowledge Drs. Tara O’Rourke and Donald Williams for their assistance with patient recruitment.

## Declaration of Interest Statement

None of the authors have any relevant financial disclosures.

## Notes

### Competing Interest Statement

The authors have declared no competing interest.

### Clinical Trial

NCT03511495

### Funding Statement

No external funding was received in relation to this project.

